# P-Values for Two-Step Genome-Wide Gene–Environment Interaction Scans

**DOI:** 10.1101/2023.06.27.23291946

**Authors:** Juan Pablo Lewinger, Bo Yuan, Eric S. Kawaguchi, W. James Gauderman

## Abstract

Two-step testing has emerged as the leading approach for genome-wide gene-environment interaction scans, providing greater power than standard single-step methods in nearly all plausible scenarios. This gain in power has enabled several recent discoveries in gene-environment interactions. However, despite controlling the genome-wide type I error rate, two-step tests typically lack valid p-values, hindering comparisons with single-step results. We demonstrate how multiple-testing adjusted p-values can be defined for two-step tests using standard multiple-testing theory, and how these can be further scaled to allow valid comparisons with single-step tests.

## 1 Introduction

Recent gene-environment (*G* × *E*) findings (Carreras-Torres et al., 2023; Virolainen et al., 2023) have been facilitated by two key developments: (1) the pooling of large sample sizes across studies with harmonized genotype and exposure data, and (2) the adoption of two-step genome-wide interaction scan (GWIS) approaches. Two-step approaches have been proposed to increase power for detecting *G* × *E* interactions while controlling the family-wise error rate across the genome for the most common outcome types, including binary (Gauderman et al., 2013; Hsu et al., 2012; Kooperberg & LeBlanc, 2008; Murcray et al., 2009, 2011; Wang et al., 2022), quantitative (Paré et al., 2010; Zhang et al., 2016), and time-to-event traits (Kawaguchi et al., 2022).

In contrast to standard single-step GWIS—which tests each SNP–exposure interaction at a stringent genome-wide significance level (e.g., *α* = 5 × 10^*−*8^; see (Dudbridge & Gusnanto, 2008))—two-step methods first use a screening step to prioritize SNPs, followed by a testing step that uses more permissive, data-adaptive significance thresholds for the most promising candidates.

This two-step strategy yields higher power under most biologically plausible *G×E* scenarios (Gauderman et al., 2013; Hsu et al., 2012; Kawaguchi et al., 2022; Kooperberg & LeBlanc, 2008; Murcray et al., 2009, 2011; Paré et al., 2010; Wang et al., 2022; Zhang et al., 2016). Multiple combinations of screening statistics and testing strategies have been proposed, including subset (Murcray et al., 2009) and weighted hypothesis testing (Ionita-Laza et al., 2007; Kawaguchi et al., 2023). However, none of these methods currently provide a principled framework for computing multiple-testing-adjusted p-values, thereby limiting interpretability and comparability across approaches.

In this note, we propose adjusted p-values for two-step GWIS procedures based on standard multiple-testing theory. Our approach is completely general and can be applied with any combination of the existing screening and testing strategies. These adjusted p-values enhance transparency and interpretability, allowing for valid comparisons across *G* × *E* methods.

## 2 Methods

Let *Y* denote the outcome (binary, continuous, or time-to-event), *E* a single environmental exposure, and *G*_1_, …, *G*_*M*_ the genotypes for *M* SNPs measured or imputed in *N* subjects. A two-step GWIS consists of:

### Step 1 (Screening)

For each SNP, a screening statistic is calculated - typically based on its marginal association with the outcome Kooperberg and LeBlanc, 2008, the exposure Murcray et al., 2009 or a combination of both Gauderman et al., 2013. The screening statistic ranks SNPs according to how promising they are for *G* × *E* interaction testing; under most scenarios, SNPs truly involved in interactions will tend to exhibit larger screening statistics.

### Step 2 (Testing)

SNPs are formally tested for interaction using a significance level *α*^*∗*^ that depends on their screening result in Step 1.

Two common strategies for implementing Step 2 are:

### Subset Testing

Only the top *m ≪ M* SNPs exceeding a predefined screening threshold are tested for interaction. A Bonferroni correction is applied using *m* as the denominator:

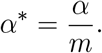

### Weighted Hypothesis Testing

All *M* SNPs are tested, but with bin-specific significance thresholds based on their Step 1 rankings. Specifically, SNPs are sorted by their screening statistic and assigned to *K* bins, *B*_1_, …, *B*_*K*_, with corresponding sizes *m*_1_, …, *m*_*K*_. The top *m*_1_ SNPs (most promising) are placed in *B*_1_, the next *m*_2_ in *B*_2_, and so on. Bin *k* = 1, …, *K* is tested at a bin-wise error rate (BWER), *α*_*k*_, where 0 *< α*_*K*_ *< … < α*_1_, so that higher-priority bins are tested at less stringent significance levels. The *α*_*k*_ values form a partition of the overall significance level *α*, such that 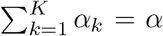. A common bin-wise partition of the *α* level and choice of bin sizes is (Ionita-Laza et al., 2007):

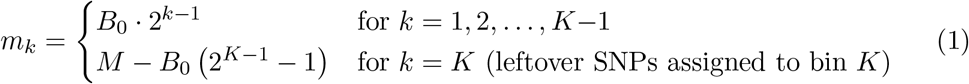

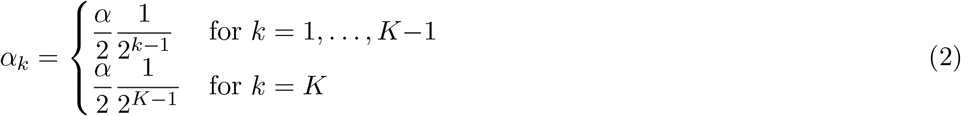

with a typical choice of *B*_0_ = 5 and where *K* = min(*k ∈* N : *B*_0_ *·* 2^*k*^ *> M* ). Within each bin, SNPs are tested using a Bonferroni correction—that is, the bin-wise error rate *α*_*k*_ is further divided by the number of SNPs assigned to the bin.

For example, with these choices for *α*_*k*_ and *m*_*k*_, the top 5 SNPs are placed in bin 1 and tested at level *α*_1_*/*5; the next 10 SNPs are placed in bin 2 and tested at *α*_2_*/*10; the next 20 SNPs go to bin 3 and are tested at *α*_3_*/*20, and so on. Overall, this scheme preserves the FWER *≤ α*, while assigning more permissive significance thresholds to SNPs considered more promising based on Step 1, thereby increasing power. Recent extensions to this strategy (Kawaguchi et al., 2023) aim to reduce power loss due to SNPs with strong marginal effects but no true interaction, and to account for correlations due to linkage disequilibrium (LD) during the testing step.

### 2.1 Multiple Testing

The key to constructing valid p-values for two-step procedures is to recognize them as special cases of multiple testing procedures. Suppose we aim to test a family of null hypotheses *H*_0_(1), …, *H*_0_(*M* ), for example, that SNP *j*, for *j* = 1, …, *M*, does not interact with exposure *E* to affect outcome *Y* . The family-wise error rate ( FWER) i s the probability of making at least one false rejection:

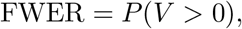

where *V* denotes the number of false positives. A FWER-controlling *multiple testing procedure* (MTP) (Dudoit & van der Laan, 2008) specifies a s et o f test s tatistics *T* _*j*_ a nd corresponding critical values 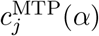 such that rejecting *H*_0_(*j*) when 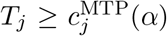 ensures FWER *≤ α*.

#### Example: Bonferroni Procedure

Let *c*_*j*_(*α*) denote the critical value for testing the individual hypothesis *H*_0_(*j*), such that *P* (*T*_*j*_ *≥ c*_*j*_(*α*) | *H*_0_(*j*)) *≤ α*. While *c*_*j*_ may vary with *j*, it is often constant across tests. Then, the Bonferroni multiple-testing critical value for *j* = 1, …, *M* is given by,

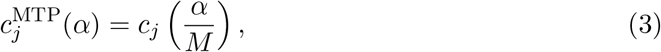

which ensures control of the FWER at level *α* regardless of test dependence.

#### Example: Two-step Subset Procedure

In this procedure, only the top *m ≪ M* SNPs—selected based on a screening statistic—are tested for interaction in Step 2. A SNP that passes the screening step is declared significant if its Step 2 p-value satisfies *p*_*j*_ *< α/m*, corresponding to a critical value:

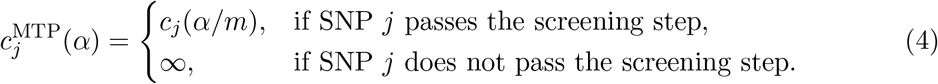

This procedure has been shown to control the FWER (Kooperberg & LeBlanc, 2008; Murcray et al., 2009), and thus defines a valid MTP in which the critical value depends on the Step 1 screening outcome.

#### Example: Two-step Weighted Bonferroni Procedure

In the weighted Bonferroni two-step procedure, the Step 2 critical value depends on the bin to which SNP *j* is assigned based on its screening statistic. Specifically, under the binning scheme defined by (2) and (1), suppose SNP *j* is assigned to bin 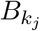, which has size 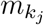 and is allocated a bin-wise error rate 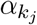 . The corresponding critical value is:

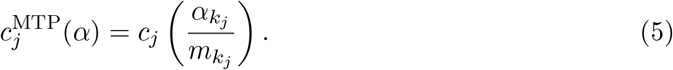

This approach also guarantees FWER control (Gauderman et al., 2013; Ionita-Laza et al., 2007), and thus defines a valid MTP in which the critical values depend on the Step 1 screening results.

### 2.2 Adjusted p-values

Since two-step procedures are special cases of multiple testing procedures, their adjusted p-values can be derived from the general framework of multiple-testing-adjusted p-values. We begin by recalling the standard definition of an *unadjusted* p-value.

Classically, the p-value is defined as the probability, under the null hypothesis, of observing a test statistic *T* at least as extreme as the observed value *t*_obs_. If the null hypothesis is *H*_0_ : *θ ∈* Ω_0_, where Ω_0_ is the subset of parameter values consistent with *H*_0_, then

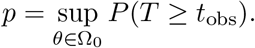

An equivalent and useful characterization (Casella & Berger, 2002) defines the p-value as the smallest significance level *α* at which the observed test statistic would lead to rejection of the null:

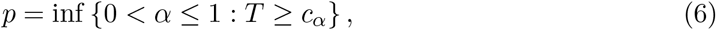

where *c*_*α*_ is the critical value corresponding to level *α*, such that type I error is controlled at *α* under *H*_0_.

This formulation extends naturally to multiple testing. Let *T*_*j*_ be the test statistic for hypothesis *H*_0_(*j*), and 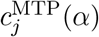 the MTP-defined critical value. Then the adjusted p-value for *H*_0_(*j*) is:

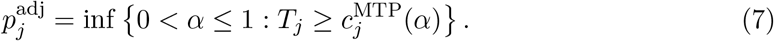

That is, 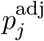 is the smallest FWER-controlled level *α* at which *H*_0_(*j*) would be rejected.

#### Example: Bonferroni Adjustment

For the classical Bonferroni procedure,

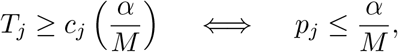

where *p*_*j*_ = inf{*α* : *T*_*j*_ *≥ c*_*j*_(*α*)} is the unadjusted p-value for *H*_0_(*j*). Thus, the adjusted p-value becomes:

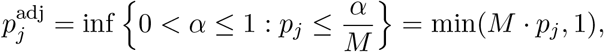

which is the familiar Bonferroni adjusted p-value.

#### Example: Two-step Subset adjustment

Based on the two-step critical values in (4) and using the general definition in (7), the adjusted p-value is:

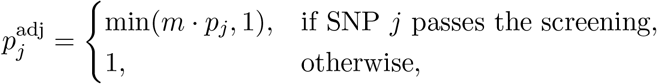

where *m* is the number of SNPs tested in Step 2.

#### Example: Two-step Weighted Bonferroni Adjustment

Under the weighted Bonferroni scheme, the Step 2 p-value is compared to 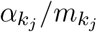. Hence, the adjusted p-value is:

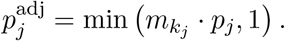

For the binning scheme defined by (2) and (1), this simplifies to:

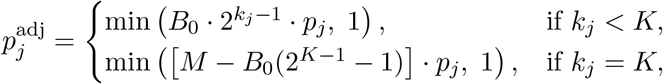

This shows that SNPs assigned to lower-index bins—ranked as more promising in Step 1—receive smaller adjusted p-values for the same Step 2 test p-value.

This framework for computing multiple-testing-adjusted p-values is general and can be applied to any two-step testing strategy. In the next section, we demonstrate its application using real data and an enhanced version of the two-step weighted Bonferroni method introduced by Kawaguchi et al. (Kawaguchi et al., 2023).

## 3 Example

To illustrate the proposed multiple-testing p-value adjustment for two-step procedures, we consider the GWIS of colorectal cancer by alcohol intake from the FIGI consortium (Jordahl et al., 2022)(Figure 1). This GWIS applied a recent extension of the weighted-Bonferroni two-step procedure (Kawaguchi et al., 2023), in which SNPs are binned based on the range of their Step 1 p-values, rather than assigning a fixed number of SNPs as in Equation 1. Specifically, SNPs are assigned to bin *B*_*k*_ based on which range their Step 1 p-value falls into. Bin 1 includes all SNPs with:

**Figure 1.**
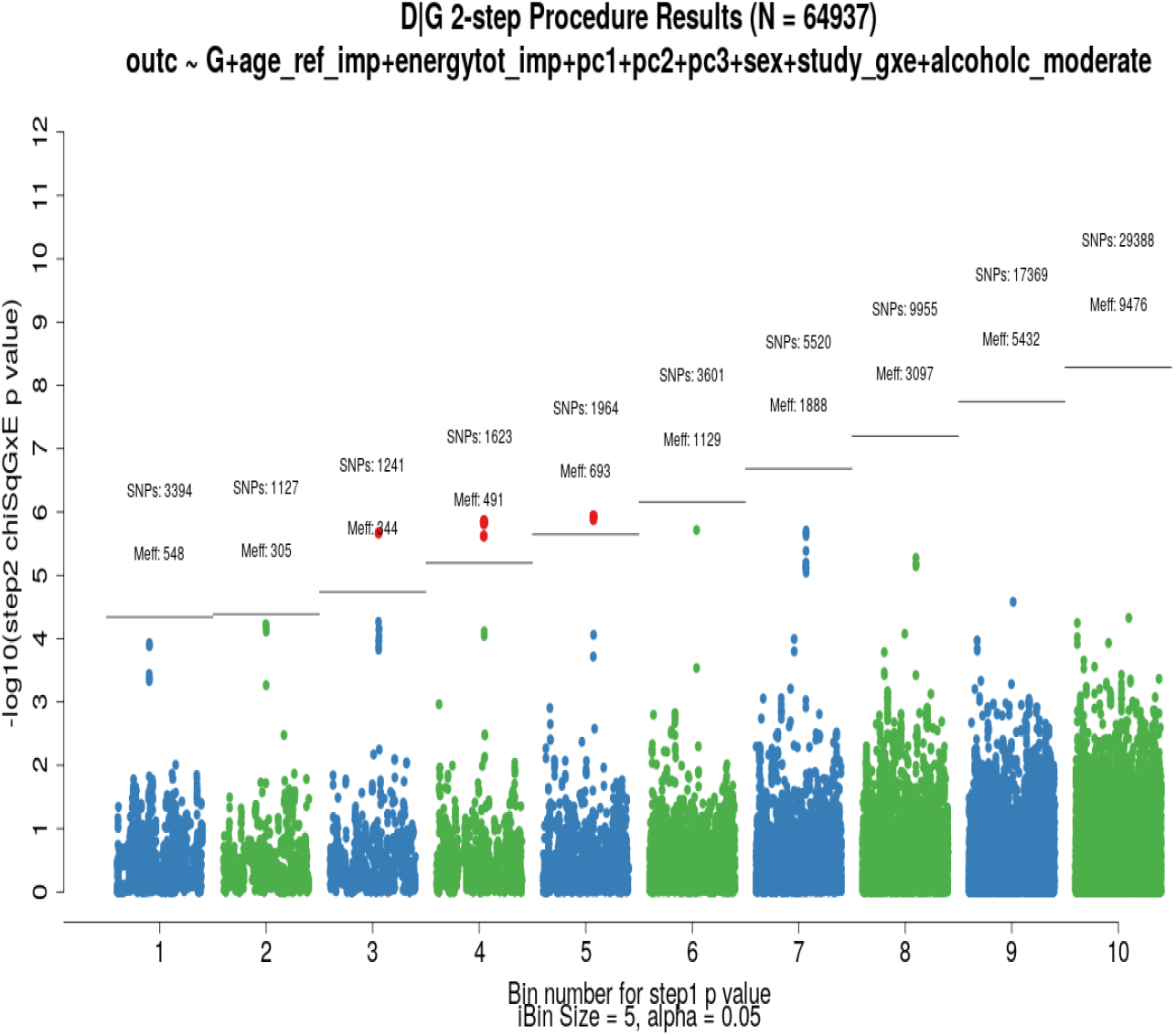
*G*× Alcohol GWIS.

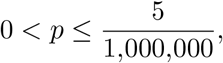

where 1,000,000 reflects the effective number of SNPs tested corresponding to a conventional genome-wide threshold of 5 × 10^*−*8^. Bin 2 includes SNPs with:

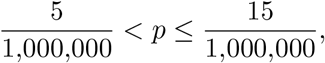

and in general, bin *B*_*k*_ includes SNPs with Step 1 p-values in the interval:

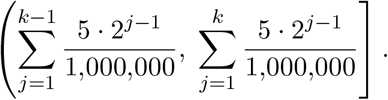

Under the null hypothesis of no marginal genetic effect and no gene–environment interaction—that is, assuming no SNP individually or in combination with alcohol affects colorectal cancer risk—the expected number of SNPs per bin is 5 in bin 1, 10 in bin 2, 20 in bin 3, and so on. In expectation, this mirrors the fixed bin assignments defined in Equation 1. A key advantage of this strategy is that it prevents “bin overcrowding” by SNPs with strong marginal effects but no interaction component (Kawaguchi et al., 2023). In addition, the bin-wise error rate (BWER) is controlled using a Bonferroni correction that accounts for the *effective* number of independent SNPs in each bin, adjusting for linkage disequilibrium. In their simulation study, Kawaguchi et al. also demonstrated improved power over the standard weighted-Bonferroni two-step procedure (Kawaguchi et al., 2023).

Figure 1 shows that one SNP in bin 3, two in bin 4, and two in bin 5 are significant (highlighted in red). For instance, the significant SNP in bin 3 has a Step 1 p-value of 3.43 × 10^*−*5^, placing it in bin 3, and a Step 2 p-value of 2.14 × 10^*−*6^. Bin 3 includes 1,421 SNPs, corresponding to 244 effectively independent tests after LD adjustment. The adjusted p-value is computed as:

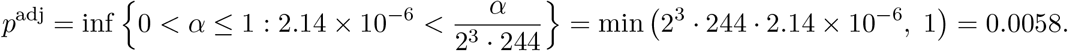

This adjusted p-value can be directly compared to a conventional significance level such as *α* = 0.05. Alternatively, dividing it by the conventional number of effective tests—assumed to be 1,000,000 SNPs, which gives rise to the genome-wide significance level of 5 ×10^*−*8^—yields 0.0058*/*1,000,000 = 5.8 × 10^*−*9^. This value can then be compared to the standard genome-wide significance threshold of 5 × 10^*−*8^.

## 4 Discussion

In this paper, we presented a method for adjusted p-values for two-step tests of *G* × *E* interactions using standard multiple-testing theory. This approach enables valid comparisons with single-step tests and offers a principled and interpretable metric for assessing statistical significance in GWIS. In addition, the availability of valid adjusted p-values can facilitate downstream meta-analyses across studies using different GWIS strategies. The two-step methods described above are all implemented in a suite of R packages. This suite includes GxEScanR (Morrison & Gauderman, 2025) to scan the genome and generate Step-1 and Step-2 statistics, and GxETest (Fu et al., 2025) to use these statistics to form 2-step tests and compute the adjusted p-values described in this paper. We anticipate that our methods and software framework will be a valuable resource for researchers conducting GWIS and will contribute to more consistent and transparent reporting of results.

## Data Availability

The data and code that support the findings of this study are available from the corresponding author upon reasonable request.

## Ethics Approval

This study used publicly available summary statistics and simulated data and did not involve human participants directly; therefore, ethics approval and consent were not required.

## Acknowledgements

The work was partially supported by NIH grants T32ES013678, P01CA196569, R01CA201407, and P30ES007048. These awards had no influence over the experimental design, data analysis or interpretation, or writing the manuscript.

